# A Real World Evaluation of the safety and immunogenicity of the Covishield vaccine, ChAdOx1 nCoV- 19 Corona Virus Vaccine (Recombinant) in Health Care Workers (HCW) in National Capital Region (NCR) of India: A preliminary report

**DOI:** 10.1101/2021.04.14.21255452

**Authors:** Sushila Kataria, Pooja Sharma, Vikas Deswal, Kuldeep Kumar, Manish Singh, Sazid Alam, Vaibhav Gupta, Padam Singh, Rashmi Phogat, Smita Sarma, Nipun Patil, Rohit Dutt, Renu Saxena, Naresh Trehan

**Affiliations:** Internal Medicine, Medanta- The Medicity, Gurugram, Haryana, India; Medanta Institute of Education and Research, Gurugram, Haryana, India; Medanta Institute of Education and Research, Gurugram, Haryana, India, and PhD Scholar GD Goenka University; Medanta- The Medicity, Gurugram, Haryana, India; Department of Microbiology, Medanta- The Medicity, Gurugram, Haryana, India; School of Medical and Allied Sciences, GD Goenka University, Gurugram, Haryana, India; Laboratory Medicine, Medanta- The Medicity, Gurugram, Haryana, India

**Keywords:** SARS-CoV-2, Covishield, ChAdOx1 nCoV- 19, real world, safety, immunogenicity vaccination, healthcare workers, early response

## Abstract

**Background:** The SARS-CoV-2 pandemic has severely impacted health systems, economic and social progress globally in 2020. The rollout of vaccines in several parts of the world is being hailed as a solution to the crisis. With newer and more virulent serotypes on the horizon and limited vaccine available, evaluation of safety and immunogenicity is critical for rationalization of vaccine use in public health.

**Objective:** To evaluate real world safety, and, immunogenicity of the Covishield vaccine, ChAdOx1 nCoV-19 Corona Virus Vaccine (Recombinant) in Health Care Workers (HCW) during the national vaccine roll out in the NCR, New Delhi. The safety is evaluated through Adverse Events and Serious Adverse Events reported though enhanced pharmacovigilance protocols, and, the immunogenicity by quantitative determination of anti-S1 and anti-S2 specific IgG antibodies to SARS-CoV-2 in serum samples collected before the receipt of the vaccine and 14 days after dose 1, using the fully automated LIAISON® SARS-CoV-2 S1/S2 IgG test using the chemiluminescence immunoassay (CLIA)

**Results:** In the two weeks after immunization with the Covishield vaccine {ChAdOx1 nCoV-19 Corona Virus Vaccine (Recombinant)}, none of the 1638 evaluated participants reported any serious adverse events (ie require hospitalization or emergency room visit). Solicited adverse events reported via daily diary cards included pain (62.7%) and soreness (24.1%) at injection site as most common, whereas fever (48.4%), headache (43.4%), myalgia (38.4%), fatigue (33.4%), joint pain (27.0%) and nausea (16.0%) were most common solicited systemic adverse events on day 1. Majority of local and systemic adverse events were seen in first 2 days post vaccination and thereafter they resolved. Lesser reactogenicity was observed in subjects with age >50 years. No major difference was observed in adverse events when subjects were stratified based on history of COVID 19 disease or baseline seropositivity. In our study serostatus improved from 48.2% positive at baseline to 79.0% positive 2 weeks following first dose of vaccination. After first dose of vaccination overall higher percentage (98.2%) of seropositivity rates were observed in those with past history of COVID 19 disease

**Conclusion:** The Covishield vaccine {ChAdOx1 nCoV-19 Corona Virus Vaccine (Recombinant)}, was safe and reported mild self limiting adverse events over 2-4 days and had an good early (within 2 weeks) seroresponse. This holds the promise of far reaching impact on vaccine availability for a larger population and thereby providing a widespread coverage.

## 1. Introduction

The first January 2020, launched the new year at the WHO offices with an IMST (Incident Management Support Team) created across the three levels of the organization, putting the organization on an emergency footing for dealing with the outbreak of a cluster of cases of pneumonia in Wuhan, Hubei, which, progressed rapidly to the declaration of a Covid-19 pandemic on March 11, 2020 **(1)**.Worldwide massive responses were put into action, including lockdowns, social distancing measures, quarantine, several drugs and technologies swung into action to minimize morbidity and mortality. Despite the best efforts of humankind, over 106,125,682 confirmed cases and 2,320,497 lives worldwide have been lost to Covid 19 disease **(2)**.The infectious nature of the disease initiated the race to develop and deploy safe and effective vaccine. There are currently more than 50 COVID-19 vaccine candidates in trials **(3)**.

On Dec 11,2020 the US Food and Drug Administration (FDA) authorized the emergency use of the mRNA vaccine BNT 162b2 to Pfizer and BioNTech, against Covid 19 in individuals 16 years or older in age, this was followed by a national vaccine roll out in the US for high risk persons **(4)**. Jan 3, 2021, the Indian drugs regulator, DCG(I) CDSCO approved the Oxford-Serum Institute’s Covid-19 Vaccine Covishield and Bharat Biotech’s Covaxin for restricted Emergency Use (**5**).

To roll out the vaccine strategy in Indian The National Expert Group on Vaccine Administration (NEGVAC) for COVID-19, was instituted on 7th August 2020 to provide guidance on prioritization of population groups, procurement and inventory management, Vaccine selection and Vaccine delivery and tracking mechanism. Draft regulatory guidelines for development of vaccines with special consideration for covid-19 vaccine were released to ensure post launch safety and surveillance with special considerations for Covid Vaccines (**6**). Post Marketing Surveillance including assessment of Adverse Events Following Immunization (AEFI) and monitoring of Adverse Events of Special Interest (AESI) ie potential risk of vaccine-associated Enhanced Respiratory Disease (**ERD**) are recommended.

On 16 January 2021, India started its national vaccination programme against the SARS-CoV-2. The drive prioritizes healthcare and frontline workers, and then those over the age of 50 or suffering from certain medical conditions.

During this national vaccination campaign we have planned an observational real world study at our hospital to evaluate the safety, immunogenicity and efficacy of COVID-19 in Health Care Workers (HCW).

## 2. Methodology

This study is a prospective cohort study evaluating the safety, immunogenicity and effectiveness of the Covid-19 vaccine given as the national vaccine roll out in health care workers. The study was approved by the Institutional Ethics Committee and was conducted in a fifteen hundred bedded tertiary care hospital in the National Capital Region of Delhi, which has treated over eight thousand hospitalized Covid-19 patients. The study was conducted in accordance with International Conference on Harmonisation–Good Clinical Practice (ICH-GCP) guidelines, the Declaration of Helsinki, and local regulatory requirements.

The hospital has 6962 HCW registered for vaccination in the national vaccine roll out. All health care personnel who consented to participate in the real world cohort for the evaluation of the Covid-19 vaccine were eligible for participation. We further excluded those who had participated in any covid prophylactics or drug trials, or had received immunoglobulins and/or convalescent plasma within the three months preceding the planned administration of the vaccine (Jan 16, 2021). Those who were ineligible to receive the vaccine due to history of hypersensitive reactions, or active Covid-19 disease were also excluded from participation (**7**).

The informed consent process followed by a baseline questionnaire was completed by a doctor. The baseline questionnaire collected information on basic socio-demographic characteristics (Age and gender), health information (comorbidities, use of any supplements including alternative medicine) History of previous Covid-19 diagnosis (with severity) was also elicited (**8**).Work related information (role in the hospital, the type of exposure at work ie low or high risk was noted (**9**).

3 ml of blood was collected at baseline (Day 0, day of vaccination) and Day 14±2 (after receipt of dose 1 of the vaccine), in Serum Separated Tube (SST, Yellow Top Vacutainer) aseptically by venepuncture. The samples were stored at centrifuged at 3000 to 3500 RPM for 10 minutes and the separated serum was kept at 2°-8°C until further testing (within a week). The participants each received a printed daily dairy to capture solicited adverse events and their severity. A project hotline (dedicated phone line) documented any unsolicited Adverse Vaccine reactions, and Vaccine Associated Adverse Events of Special Interest (AESI), such as Vaccine Associated Enhanced Respiratory Disease (ERD) and any report of symptomatic COVID-19. Each sample, baseline questionnaire and daily diary were barcoded, and linked. The severity of adverse events was graded according to VigiFlow® for Adverse Events.

### Enhanced pharmacovigilance protocol

The research Adverse Event (AE) monitoring teams conducted an enhanced pharmacovigilance as follows

1. **Daily diary**: the participants were explained the filling of the daily diary by the doctors at the baseline visit and reminders were send on phone numbers once in the week to remind them of filling the same. The diary was available both in English and Hindi and drop boxes were created to facilitate receipt after completion and a mop up drive was organized to bring in any missing diaries.
2. **Monitoring the emergency and medicine OPD units**: The research teams monitored the participant visits to the medicine department OPD (noted as unsolicited Adverse Events)
3. **Tallying of AE reports with** those reported on PVPI (National Pharmacovigilance Program of India) by the hospital pharmacovigilance team

### Outcomes

The primary endpoint was to assess the safety of the vaccines by actively reported AE/SAE in the first week of vaccination after each dose of the vaccine, and to assess the safety of the vaccine by occurrence of unsolicited Adverse Vaccine reactions, Vaccine associated Adverse Events of Special Interest (AESI), such as Vaccine Associated Enhanced Respiratory Disease (ERD) after both doses of vaccination [Time Frame: from Day 0 till End of study at Day 118±7 days].

Baseline sero-prevalence of SARS CoV-2 S1/S2 IgG (Baseline, D-6 to 0), humoral immunogenicity (Seroconversion and antibody quantification) of COVID-19 vaccination [at Day -6/0, Day 14±2 days, Day 56±7 days, Day 118±10 days], study of the incidence of symptomatic COVID-19 infection among vaccinated healthcare workers and its correlation with antibody levels. [Time Frame: 118±7 days post vaccination] and assessment of efficacy of the COVID-19 vaccination against severe and non-severe COVID-19 [Time Frame: Day 118±7 days] are secondary endpoints.

### 2.1. Laboratory Methods

The serum was tested for the quantitative determination of anti-S1 and anti-S2 specific IgG antibodies to SARS-CoV-2 in the fully automated LIAISON® SARS-CoV-2 S1/S2 IgG by Chemiluminescence immunoassay (CLIA) technology. The analyzer automatically calculated SARS-CoV-2 S1/S2 IgG antibody concentrations expressed as arbitrary units (AU/mL) and graded the results. Test results were reported quantitatively as positive and negative. Assay ranges from 3.8 to 400 AU/mL SARS CoV-2 S1/S2 IgG. < 15 AU/mL were **reported negative** and those test results ≥15.0 AU/mL were **reported as positive**.

### 2.2. Lab Quality Assurance

The fully automated LIAISON® has been calibrated and validated according to the laboratory SOP, which is 3 samples (high, medium and low value) were run for 5 times a day for 5 consecutive days. Their mean, SD and CV% were calculated, which were within normal limits. One positive and one negative kit controls were processed daily for internal quality control. The instrument was calibrated once daily before processing the samples and also for every new lot kit used. Inter-Lab Comparison (ILC) and Repeat and split testing were conducted as per standard laboratory quality assurance protocol.

#### Procedures

The blood samples (3ml/sample) for the Immunogenicity evaluation were collected at the following time points. Pre-dose blood sample (Day - 6 to 0) was collected within 6 days prior to vaccine administration and post vaccination blood samples of all participants were collected at Day 14±2 days. Within 30 minutes, the blood samples were centrifuged at 3000 RPM for 10 minutes and immediately kept at -20°C until analysis.

##### Interpretation of results

The SARS-CoV-2 S1/S2 IgG antibody concentrations was automatically calculated using analyzer and expressed as arbitrary units (AU/mL). Test results were reported quantitatively as **positive** (≥15 AU/mL), **equivocal or negative** (<15 AU/mL). Assay ranges from 3.8 to 400 AU/mL SARS CoV-2 S1/S2 IgG was defined. The immunogenicity of the personnel was kept confidential by using a subject identification research code.

### 2.3. Data Analysis

#### Data Collection and Quality Assurance

At baseline, eligibility and medical history was assessed and informed consent was taken from all participants. For data capture each participant was assigned a unique study code, which facilitated linking of participant data with barcode of sero sample. The data were entered into e-HIS (electronic health information system) template which maintains an audit trail and was then exported into Excel sheets. Quality Assurance of the data was undertaken by an independent QA coordinator. **Safety assessment cohort** included all participants who received at least one dose of the vaccine and provided information on local and systemic adverse events. Severity of adverse events are graded as mild, moderate and severe from the patient reported daily diary. **Immunogenicity cohort** included all participants who received at least one dose of the vaccine and presented with two samples for immunogenicity evaluation (Day-6 to 0) (T1) and Day 14±2 (T2).

#### Statistical Methods

The profile of study participants had been presented according to role in the hospital, nature of exposure, demographics (age and gender) and history of COVID-19 disease. Detailed analysis of SARS CoV-2 S1/S2 IgG titers has been undertaken in terms of median (IQR) and sero positivity (defined as SARS CoV-2 S1/S2 IgG titers ≥15 AU/mL) with 95% confidence interval. Sero positivity was further analyzed according to participants’ role in the hospital, nature of exposure, age, gender and history of Covid-19 disease with severity of disease and duration. The differences in SARS CoV-2 S1/S2 IgG levels and sero positivity at T1 (Day-6 to 0) and T2 (14±2) by baseline characteristics has been studied. An analysis of local and systemic solicited adverse events has been undertaken according to their levels and severity. Further detailed analysis of commonly reported SAE’s (> 10%) has been undertaken with baseline characteristics. Appropriate tests of significance (Z test, Paired t test and two sample independent t test) have been used to derive the inferences. All analysis was done using SPSS software, version 24.0. p - value < 0.05 has been considered statistically significant. MedDRA (https://www.meddra.org) has been used to code for adverse events. Coding was done with preferred term (PT) and lowest level term (LLT) (**10**).

##### Study Flow diagram

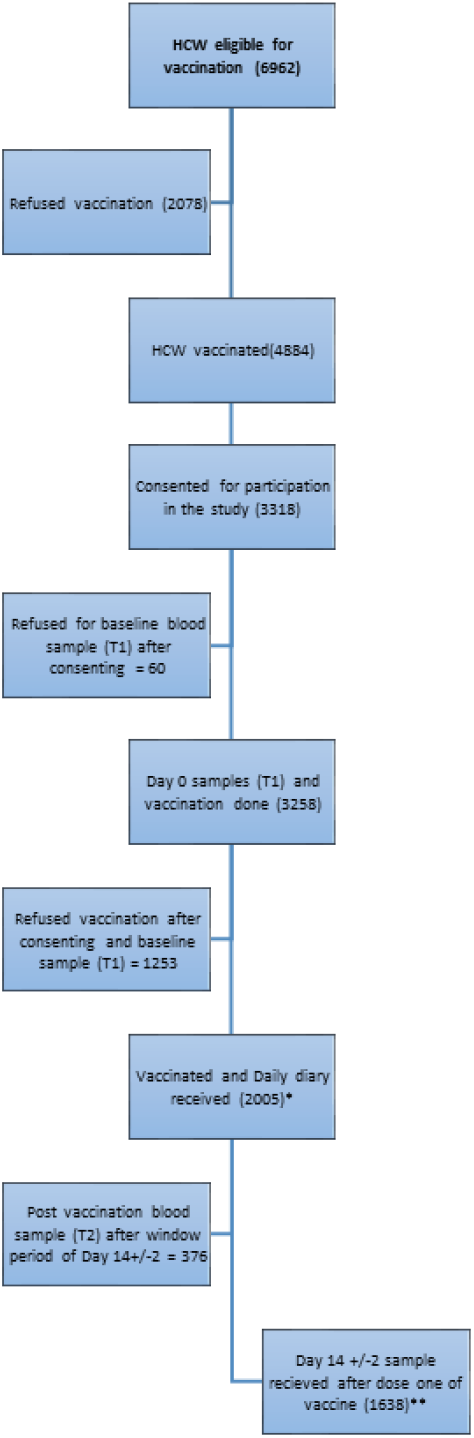

*Safety cohort

**Immunogenicity cohort

In all 6962 Healthcare Workers at our hospital were eligible to receive the covid vaccine during the Government vaccination drive. Of these refused or were not available for vaccination and received the vaccine. Out of these 3318 consented to participation in the study, 3258 gave the sample at baseline (T1) and 1638 at Day 14±2 (T2) (immunogenicity cohort),.The solicited adverse events by daily diary were collected from 2005 (safety cohort).

## 3.0 Results and Discussion

**Table 1:** Baseline Characteristics.

|  | Number of<br>Participants<br>(n=1638) | Percent<br>(%) |
| --- | --- | --- |
| Gender |  |  |
| Male | 1101 | 67.2% |
| Female | 537 | 32.8% |
| Age (Years) |  |  |
| ≤ 50 | 1509 | 92.1% |
| > 50 | 129 | 7.9% |
| Median (IQR) | 32 (26 – 40) |  |
| Role in the hospital |  |  |
| Clinical HCW | 535 | 32.7% |
| Non Clinical HCW | 1103 | 67.3% |
| Nature of Exposure |  |  |
| Low Risk Exposure | 1306 | 79.7% |
| High Risk Exposure | 332 | 20.3% |
| History of COVID-19 Disease |  |  |
| No | 1419 | 86.6% |
| Yes | 219 | 13.4% |
| Severity of COVID-19 (n=219) |  |  |
| Mild | 205 | 12.5% |
| Moderate | 10 | 0.6% |
| Severe | 4 | 0.2% |
| History of COVID-19 Disease (Duration) (n=219) |  |  |
| < 3 months | 87 | 5.3% |
| > 3 months | 132 | 8.1% |

The total number of participants in the study were 1638 **Immunogenicity cohort** with due representation of both male and female genders; clinical and non-clinical roles, low and high risk of exposure, history of COVID-19. This number was adequate to undertake disaggregated analysis on outcome parameters. Importantly **Safety assessment cohort consisted of 2005 participants**.

**Table 2:** Change in Sero positivity and median SARS CoV-2 S1/S2 IgG titres at T1 (Day -6 to 0 - Baseline) & T2 (Day 14±2 after 1^st^ dose of vaccination)

| **Parameter** | **T1**<br>**(n/N = 789/1638)** | **T2**<br>**(n/N = 1294/1638)** |
| --- | --- | --- |
| **SARS CoV-2 S1/S2 IgG Sero Positivity ($\geq 15$ AU/ml)**<br>**n (%) with 95% C. I.** | 48.2 (45.7 – 50.6) | 79.0 (76.9 – 80.9) |
| **SARS CoV-2 S1/S2 IgG titres among seropositive subjects**<br>**Median (IQR)** | 42.4 (27.3 – 76.2) | 367.5 (112.7 – 400) |

**n – Participants with SARS CoV-2 S1/S2 IgG $\geq 15$ AU/ml**

**Table 3:**
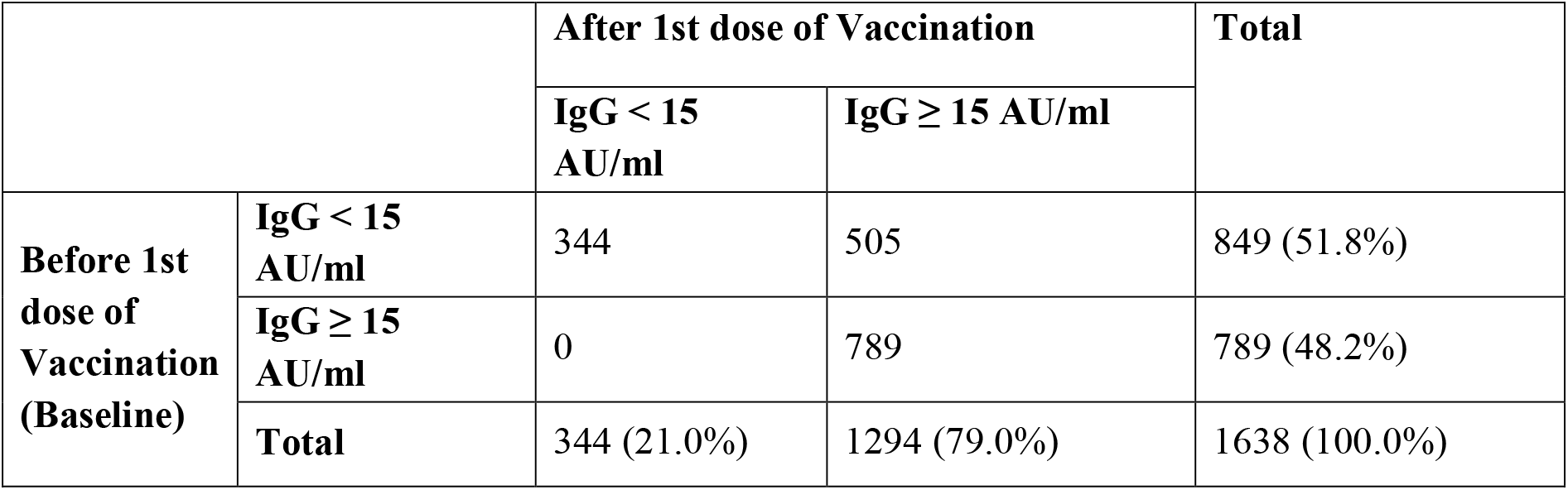
Improvement in Sero Positivity (SARS CoV-2 S1/S2 IgG titres) after 1st dose of Vaccination (n=1638)

It is observed that 51.8 % baseline seronegative turned seropositive whereas none among baseline seropositive slipped below 15 AU/ml (ie turn seronegative)

**Table 4:** Diametric shift in SARS CoV-2 S1/S2 IgG titres after 1^st^ dose of vaccination (n=1638)

| **IgG Levels** | **Before 1st dose of Vaccination (Baseline) (n=1638)** | **After 1st dose of Vaccination (n=1638)** |
| --- | --- | --- |
| **≤ 3.8** | 542 (33.1%) | 142 (8.7%) |
| **3.9 - 15.0** | 307 (18.7%) | 204 (12.5%) |
| **15.1 - 50.0** | 460 (28.1%) | 208 (12.7%) |
| **50.1 - 100.0** | 231 (14.1%) | 94 (5.7%) |
| **100.1 - 400.0** | 97 (5.9%) | 402 (24.5%) |
| **> 400** | 1 (0.1%) | 588 (35.9%) |

It is important to note that before the first dose of vaccine about one third had SARS CoV-2 S1/S2 IgG titres of 3.8 AU/ml (minimal levels) and after the first dose of the vaccine the about 36 % transitioned rapidly to SARS CoV-2 S1/S2 IgG titres of more than 400 (maximal tested by the kit) indicating a 180 degree change (mirror shift).

**Graph 1:**
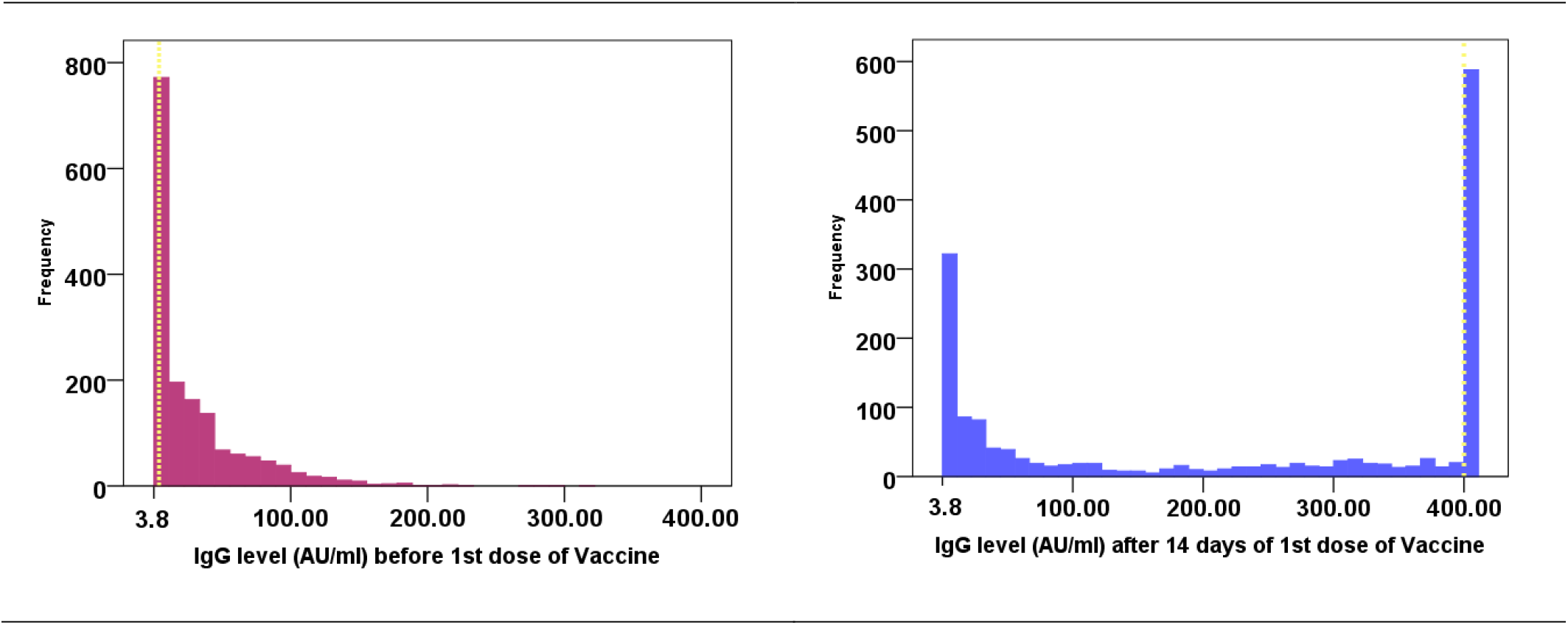
Mirror shift of IgG titres at day 14 post dose one of vaccine.

A comparison of IgG value before and after vaccination shows the shape in the curve from left skewed to right skewed.

**Table 5:** Change in SARS CoV-2 S1/S2 IgG titres before and after 1st dose of Vaccination (n=1638)

| Parameter | Before 1st dose of Vaccination (Baseline) AU/ml | After 1st dose of Vaccination AU/ml | p-value |
| --- | --- | --- | --- |
| **Participants with IgG < 15 before 1<sup>st</sup> dose of Vaccination (Baseline) (n=849)** |  |  |  |
| **Mean±SD (Range)** | 5.4±2.9 (3.8 - 14.8) | 94.6±135.2 (3.8 - 400) | < 0.0001* |
| **Median (IQR)** | 3.8 (3.8 - 6) | 24.4 (5.5 - 106.5) | < 0.0001* |
| **Participants with IgG ≥ 15 before 1<sup>st</sup> dose of Vaccination (Baseline) (n=789)** |  |  |  |
| **Mean±SD (Range)** | 56.9±42.7 (15.2 - 400) | 351±87.8 (19.1 - 400) | < 0.0001* |
| **Median (IQR)** | 42.4 (27.3 - 76.2) | 400 (329.5 - 400) | < 0.0001* |
| **All Participants (n=1638)** |  |  |  |
| **Mean±SD (Range)** | 30.2±39.3 (3.8 - 400) | 218.1±172 (3.8 - 400) | < 0.0001* |
| **Median (IQR)** | 13.6 (3.8 - 41.1) | 257 (22.5 - 400) | < 0.0001* |

\*P value <0.05, statistically significant

The change in SARS CoV-2 S1/S2 IgG titres before and after first vaccination are highly significant both in terms of mean and median. Further the change is higher for those who had SARS CoV-2 S1/S2 IgG titres ≥ 15 AU/ml at baseline.

**Table 6:** Covid-19 seropositivity analyzed by baseline characteristics at T1 and T2.

|  | n =<br>1638 | Covid-19 Sero Positivity (IgG ≥15.0 AU/ml) |  | Δ =<br>T1 –<br>T2 | p-value |
| --- | --- | --- | --- | --- | --- |
|  |  | Before 1st dose of Vaccination<br>(Baseline) (T1)<br>n (%) (95%CI) | After 1st dose of Vaccination<br>(T2)<br>n (%) (95%CI) |  |  |
| Gender |  |  |  |  |  |
| Male | 1101 | 533 (48.4%) (45.4 - 51.4)% | 864 (78.5%) (75.9 - 80.9)% | 30.1 | 0.344 |
| Female | 537 | 256 (47.7%) (43.4 - 52.0)% | 430 (80.1%) (76.4 - 83.4)% | 32.4 |  |
| Age (Years) |  |  |  |  |  |
| ≤ 50 | 1509 | 731 (48.4%) (45.9 - 51.0)% | 1210 (80.2%) (78.1 - 82.2)% | 31.8 | 0.006* |
| > 50 | 129 | 58 (45.0%) (36.2 - 54.0)% | 84 (65.1%) (56.2 - 73.3)% | 20.1 |  |
| Role in the hospital |  |  |  |  |  |
| Clinical HCW | 535 | 218 (40.7%) (26.5 - 45.0)% | 418 (78.1%) (74.4 - 81.6)% | 37.4 | <<br>0.0001* |
| Non Clinical HCW | 1103 | 571 (51.8%) (48.7 - 54.7)% | 876 (79.4%) (76.9 - 81.8)% | 27.6 |  |
| Nature of Exposure |  |  |  |  |  |
| Low Risk Exposure | 1306 | 631 (48.3%) (45.6 - 51.1)% | 1042 (79.8%) (77.5 - 81.9)% | 31.5 | 0.259 |
| High Risk Exposure | 332 | 158 (47.6%) (42.1 - 53.1)% | 252 (75.9%) (70.9 - 80.4)% | 28.3 |  |
| History of COVID-19 Disease |  |  |  |  |  |
| No | 1419 | 600 (42.3%) (39.7 - 44.9)% | 1079 (76.0%) (73.7 - 78.2)% | 33.7 | <<br>0.0001* |
| Yes | 219 | 189 (86.3%) (81.0 - 90.6)% | 215 (98.2%) (95.4 - 99.5)% | 11.9 |  |
| Severity of COVID-19 (N=219) |  |  |  |  |  |
| Mild | 205 | 177 (86.3%) (80.9 – 90.7) % | 202 (98.5%) ( 95.8 – 99.7) % | 12.2% | 0.807 |
| Moderate | 10 | 8 (80%) (44.4 – 97.5) % | 9 (90%) (55.5 – 99.7) % | 10.0 % |  |
| Severe | 4 | 4 (100%) (39.8 – 100.0) % | 4 (100%) (39.8 – 100.0) % | 0.0% |  |
| History of COVID-19 Disease (time since onset) N=219 |  |  |  |  |  |
| < 3 months | 87 | 79 (90.8%) (82.7 – 95.9) % | 84 (96.6%) (90.2 – 99.3) % | 5.8% | 0.024* |
| > 3 months | 132 | 110 (83.3%) (75.9 – 89.2) % | 131 (99.2%) (95.8 – 100.0) % | 15.9% |  |

**C. I. – Confidence Interval; p – value $< 0.05$ , statistically significant**

A subgroup analysis indicates that the change in SARS CoV-2 S1/S2 IgG titres are significantly higher for age less than 50 years and clinical health workers. The differences according to gender and level of exposure were not statistically significant. As to the differences according to history of covid, though increase is higher for those with no history, yet nothing could be concluded as the levels were already very high before vaccination.

Evidently among those with history of covid disease 86% moved from SARS CoV-2 S1/S2 IgG titres of less than 15 to more than 15. The corresponding improvement among those with no history of covid was 42% i.e. less than half of the other group.

**Graph 2:**
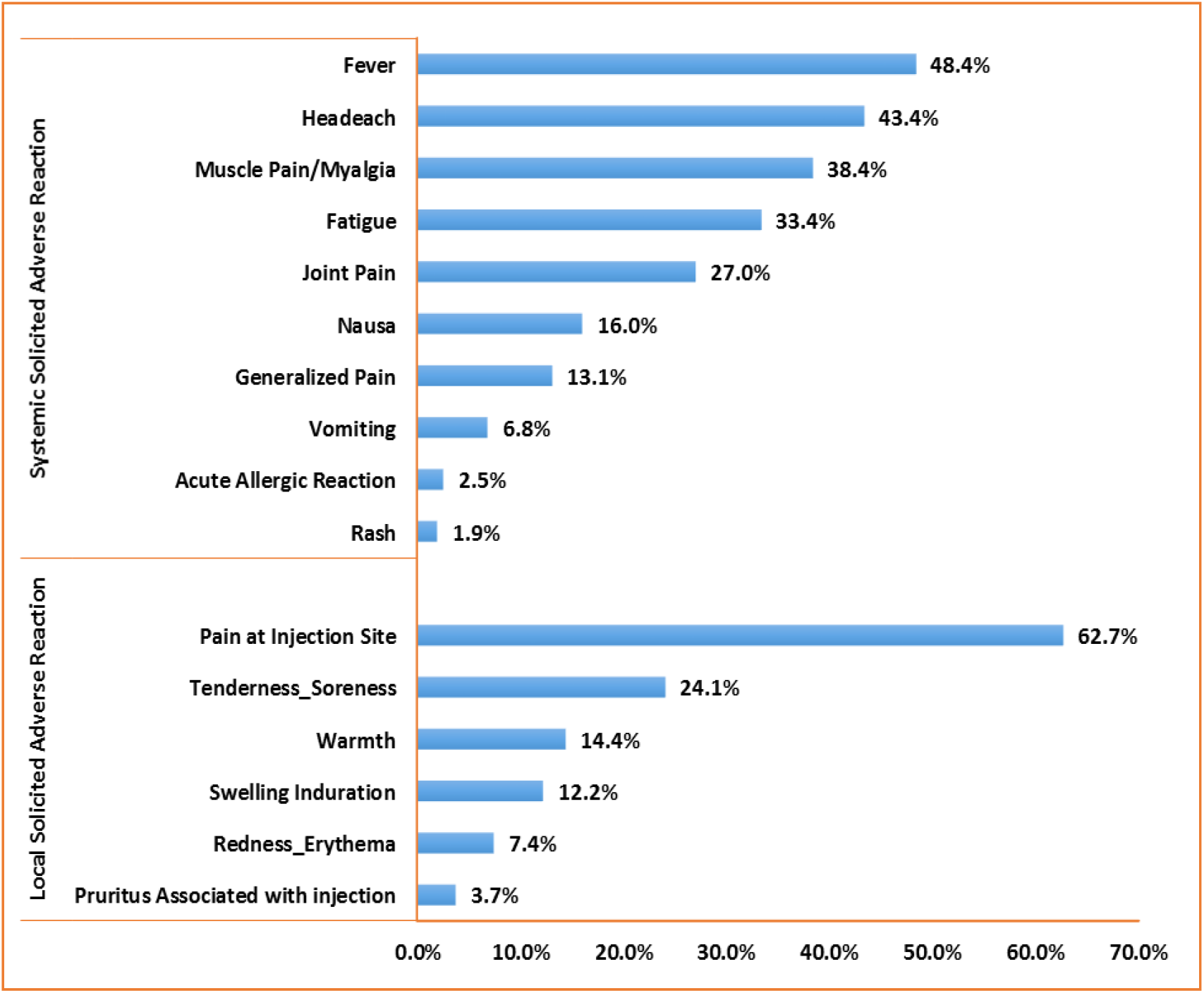
Reporting of Solicited Adverse Events (Local and Systemic)

As to the Local Adverse Events, the highest percentage reported was ***pain at injection site*** (62.7%) followed by ***Tenderness soreness*** (24.1%), ***warmth*** (14.4%) and ***swelling induration*** (12.2%). Other Symptoms reported were by less than 10% participants.

Among systemic Adverse Events the highest reported was ***Fever*** (48.4%followed by ***Headache*** (43.4%), ***Muscle pain*** (38.4%), ***Fatigue*** (33.4%), ***Joint pain*** (27.0%), ***Nausea*** (16.0%) and ***Generalized Pain*** (13.1%).Other symptoms reported were by less than 10% participants.

**Table 9:** Common Adverse Event (Local Solicited & Systemic Solicited) by past History of COVID 19 (n=1638)

|  | Before 1st dose of<br>Vaccination (Baseline) |  | Total<br>(n=1638) | p - value |
| --- | --- | --- | --- | --- |
|  | History of<br>COVID-19<br>(n=219) | No History<br>of COVID-19<br>(n=1419) |  |  |
| Local Solicited Adverse Reaction |  |  |  |  |
| Warmth | 15 (6.8%) | 154 (10.9%) | 169 (10.3%) | 0.070 |
| Pain at Injection Site | 139 (63.5%) | 701 (49.4%) | 840 (51.3%) | 0.0001* |
| Tenderness Soreness | 50 (22.8%) | 263 (18.5%) | 313 (19.1%) | 0.132 |
| Swelling Induration | 26 (11.9%) | 129 (9.1%) | 155 (9.5%) | 0.191 |
| Systemic Solicited Adverse Reaction |  |  |  |  |
| Generalized Pain | 15 (6.8%) | 123 (8.7%) | 138 (8.4%) | 0.367 |
| Fever | 85 (38.8%) | 557 (39.3%) | 642 (39.2%) | 0.901 |
| Nausea | 30 (13.7%) | 173 (12.2%) | 203 (12.4%) | 0.529 |
| Headache | 82 (37.4%) | 496 (35%) | 578 (35.3%) | 0.473 |
| Fatigue | 68 (31.1%) | 377 (26.6%) | 445 (27.2%) | 0.165 |
| Muscle Pain/Myalgia | 90 (41.1%) | 418 (29.5%) | 508 (31%) | 0.001* |
| Joint Pain | 64 (29.2%) | 305 (21.5%) | 369 (22.5%) | 0.011* |

Among commonly reported Adverse events (>10% frequency) ***Pain at Injection Site, Muscle Pain/Myalgia*** *and* ***Joint Pain*** were higher among those with history of COVID-19 as compared to those with no history of COVID-19.

**Table 10:** Common Adverse Event (Local Solicited & Systemic Solicited) by Age (n=1638)

|  | Age (Years) |  | Total<br>(n=1638) | p - value |
| --- | --- | --- | --- | --- |
|  | ≤ 50<br>(n=1509) | > 50<br>(n=129) |  |  |
| Local Solicited Adverse Reaction |  |  |  |  |
| Warmth | 164 (10.9%) | 5 (3.9%) | 169 (10.3%) | 0.012* |
| Pain at Injection Site | 785 (52%) | 55 (42.6%) | 840 (51.3%) | 0.041* |
| Tenderness Soreness | 286 (19%) | 27 (20.9%) | 313 (19.1%) | 0.584 |
| Swelling Induration | 146 (9.7%) | 9 (7%) | 155 (9.5%) | 0.315 |
| Systemic Solicited Adverse Reaction |  |  |  |  |
| Generalized Pain | 129 (8.5%) | 9 (7%) | 138 (8.4%) | 0.537 |
| Fever | 617 (40.9%) | 25 (19.4%) | 642 (39.2%) | 0.0001* |
| Nausea | 193 (12.8%) | 10 (7.8%) | 203 (12.4%) | 0.096 |
| Headache | 555 (36.8%) | 23 (17.8%) | 578 (35.3%) | 0.0001* |
| Fatigue | 423 (28%) | 22 (17.1%) | 445 (27.2%) | 0.007* |
| Muscle Pain/Myalgia | 479 (31.7%) | 29 (22.5%) | 508 (31%) | 0.029* |
| Joint Pain | 356 (23.6%) | 13 (10.1%) | 369 (22.5%) | 0.0001* |

The analysis according to age reveals significantly higher levels among those in age group less than 50 years in comparison to those in age group more than 50 years specifically local Adverse Events of ***warmth and pain***; systemic Adverse Events of ***fever*, nausea *headache, fatigue, muscle pain and joint pain***.

**Table 11:**
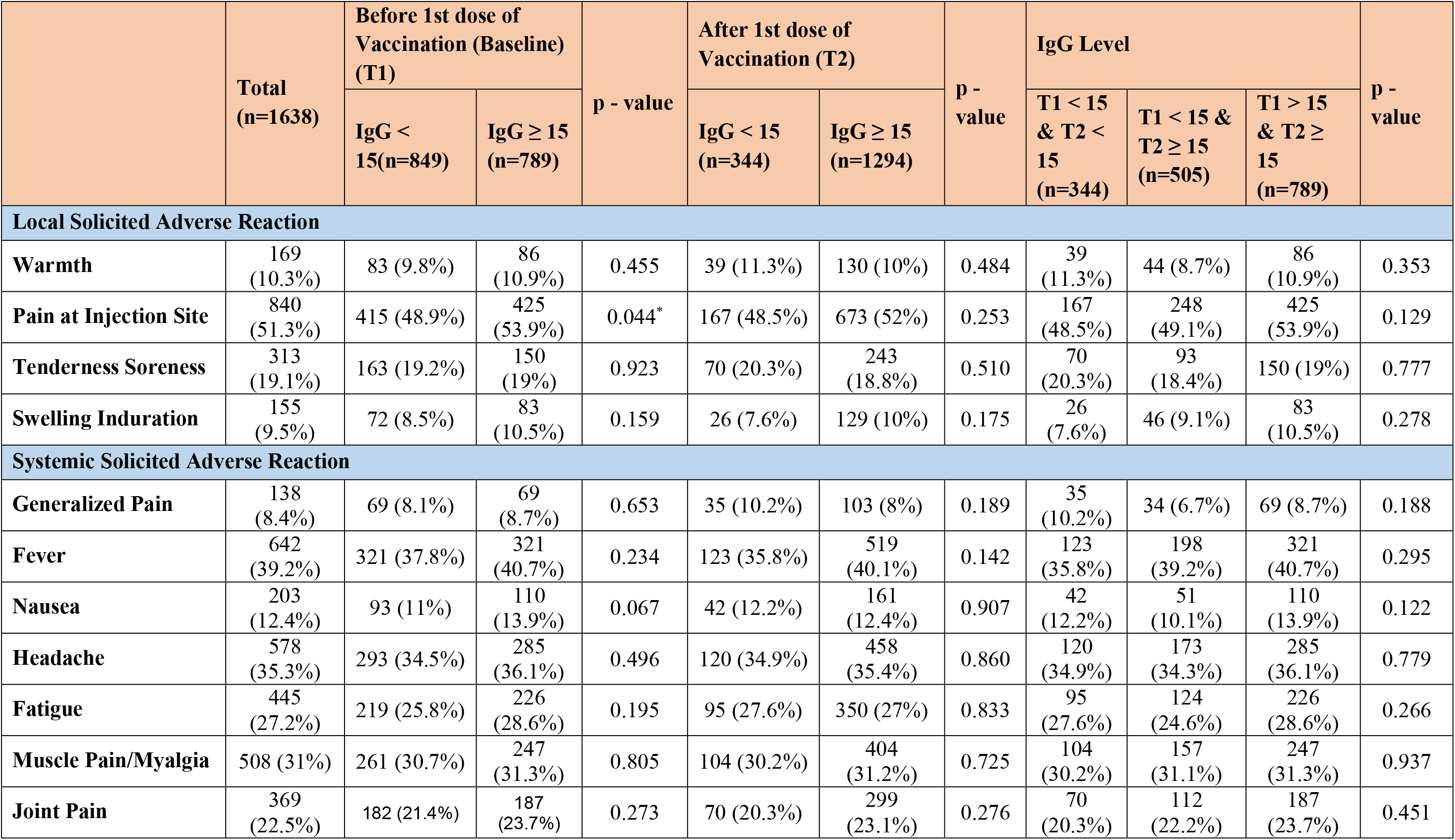
Common Adverse events according to cut-off value of IgG.

None of the observed values of common adverse events have any difference by the SARS CoV-2 S1/S2 IgG titres cut off value of 15 AU/ml

**Graph 3:**
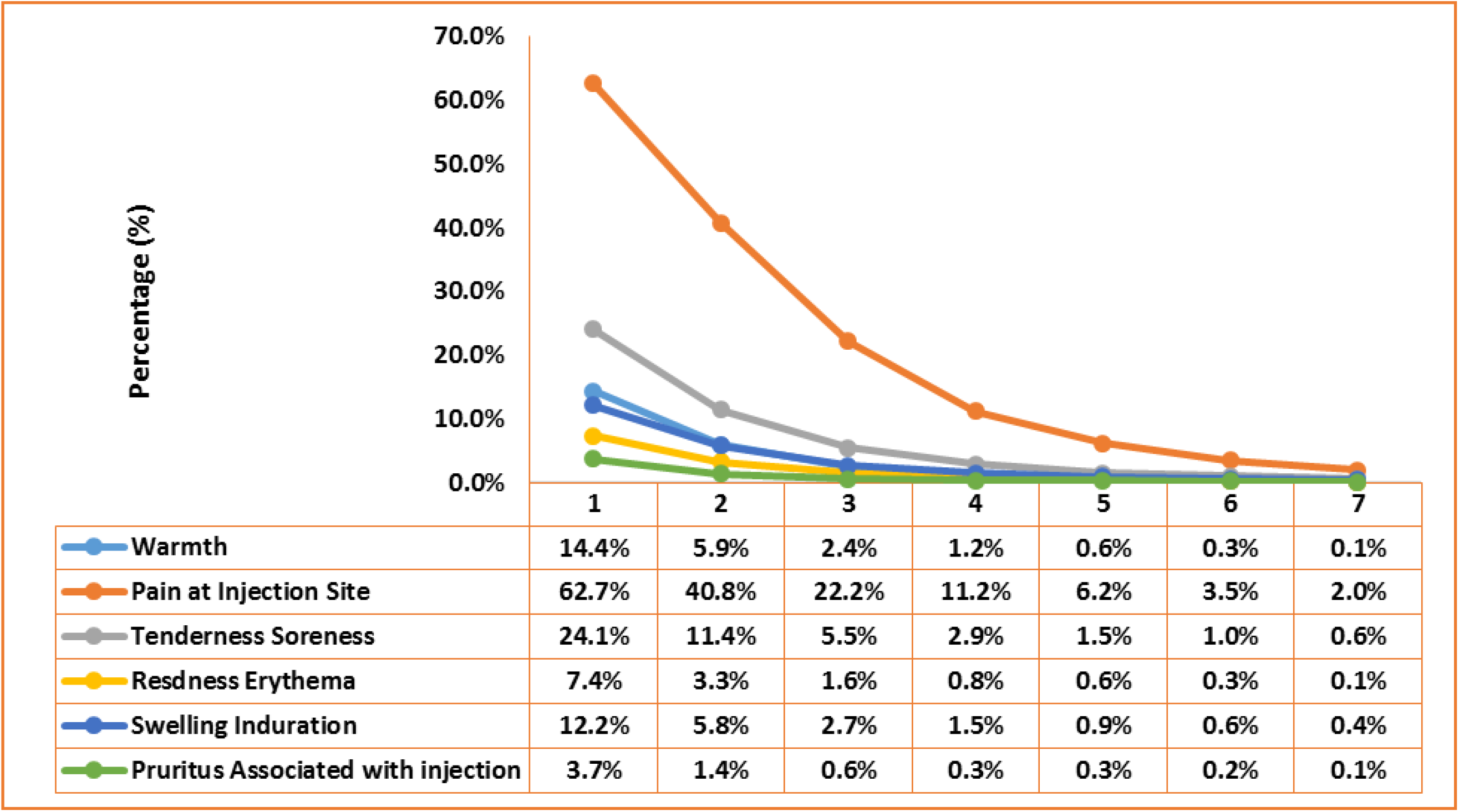
Local Solicited Adverse Events.

The local adverse events reported lasted on an average for two days. There has been steep decline from day 1 to day 4 and thereafter coming to normalcy.

**Graph 4 :**
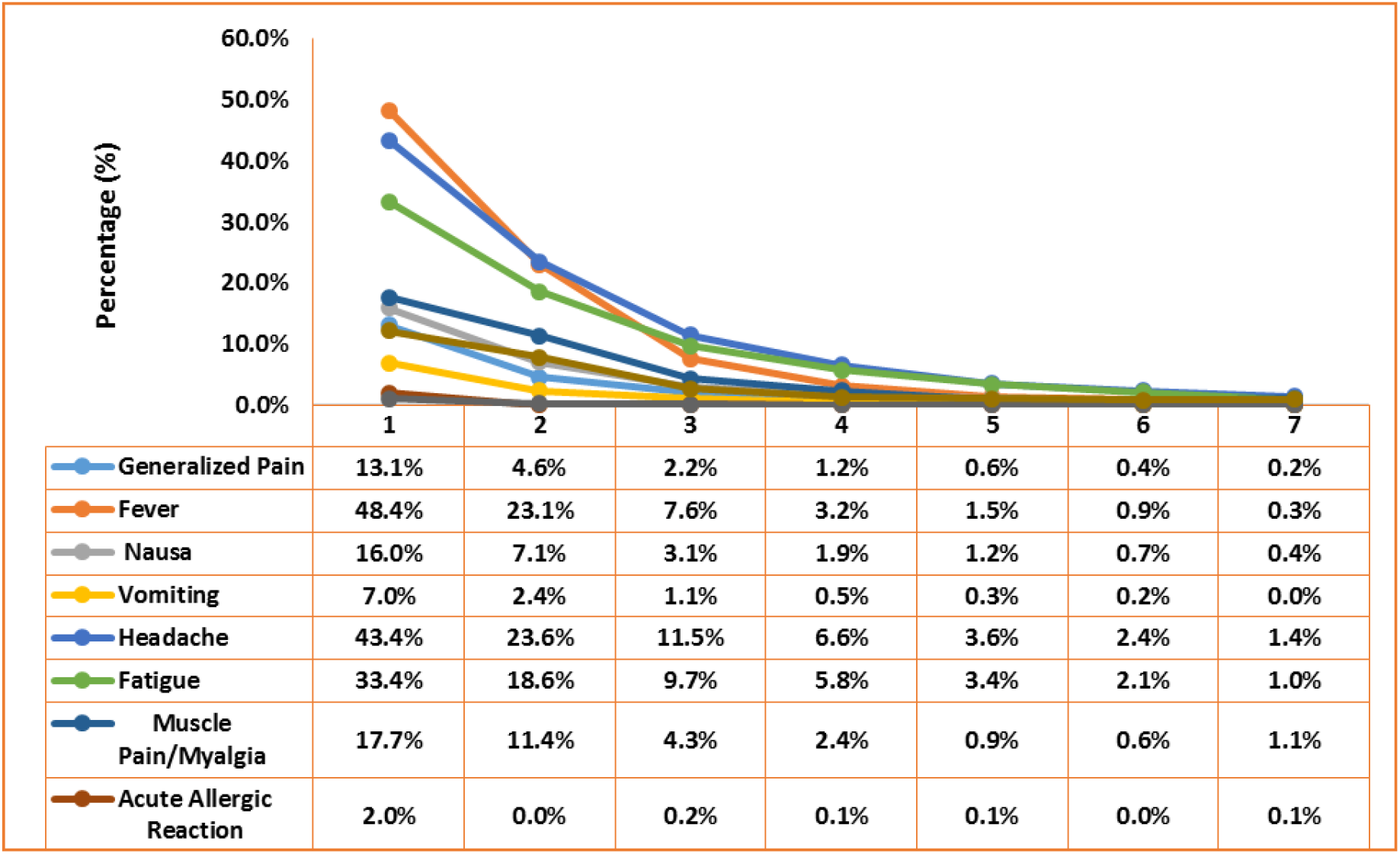
Systemic Solicited Adverse Events.

The systemic SAEs reported also lasted on an average for two days, as there has been steep decline from day 1 to day 3 and thereafter coming to normalcy.

**Graph 5:**
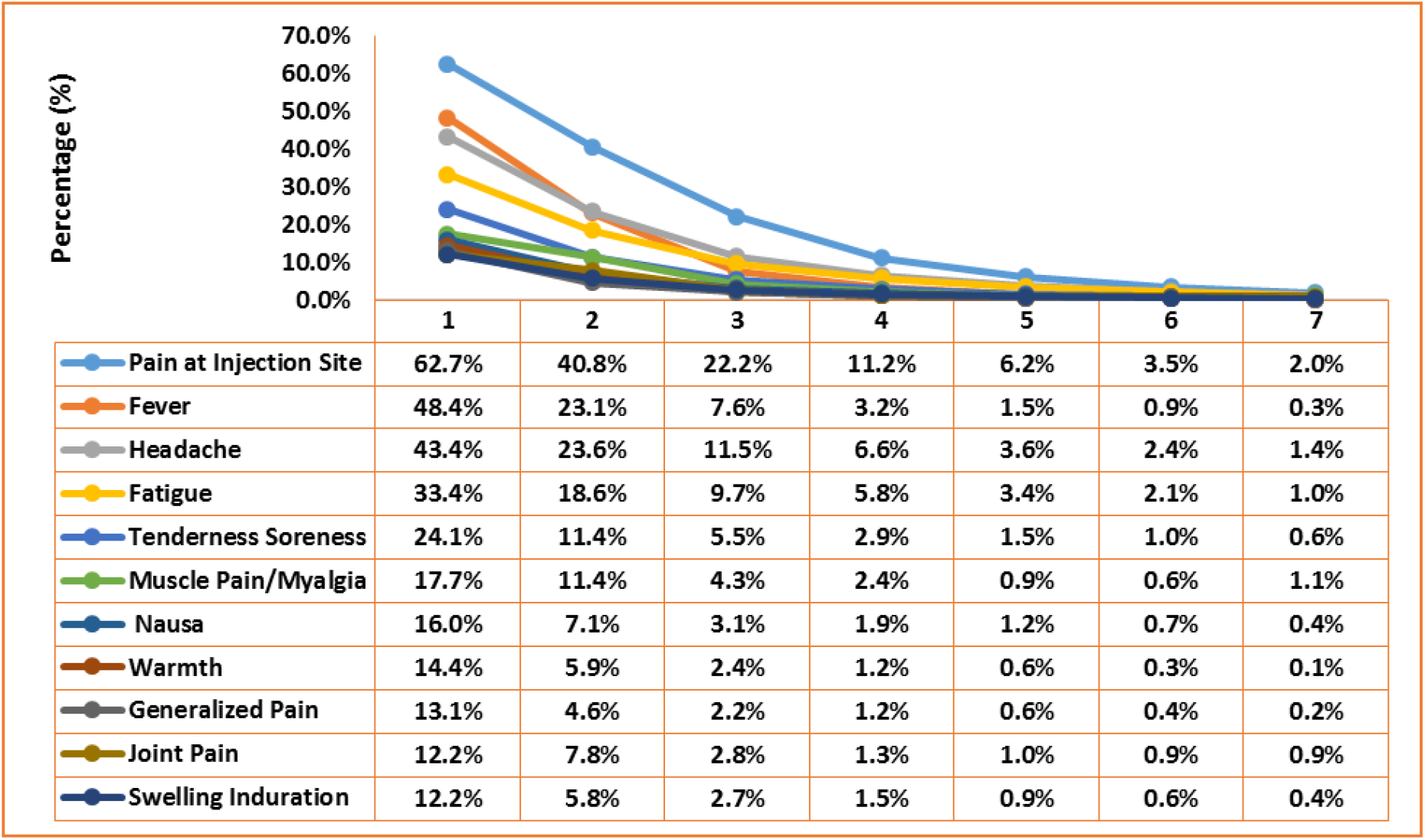
Commonly reported (>10%) Adverse Events.

For commonly reported (> 10%) Adverse Events the median duration was 3 days and there after reaching normalcy.

Importantly no case of unsolicited or Serious Adverse Events were reported in the first 14 days after vaccination.

**Table 12:** Severity of commonly reported Adverse Reactions.

| Adverse Reactions | Severity of Adverse Reactions |  |  |  |
| --- | --- | --- | --- | --- |
|  | No Reaction | Mild Reaction | Moderate Reaction | Severe Reaction |
| **Local Solicited Adverse Reaction** |  |  |  |  |
| **Warmth** | 1716 (85.6%) | 202 (10.1%) | 63 (3.1%) | 24 (1.2%) |
| **Pain at Injection Site** | 748 (37.3%) | 836 (41.7%) | 319 (15.9%) | 102 (5.1%) |
| **Tenderness Soreness** | 1522 (75.9%) | 334 (16.7%) | 110 (5.5%) | 39 (1.9%) |
| **Swelling Induration** | 1760 (87.8%) | 184 (9.2%) | 44 (2.2%) | 17 (0.8%) |
| **Systemic Solicited Adverse Reaction** |  |  |  |  |
| **Generalized Pain** | 1742 (86.9%) | 160 (8%) | 76 (3.8%) | 27 (1.3%) |
| **Fever** | 1034 (51.6%) | 577 (28.8%) | 250 (12.5%) | 144 (7.2%) |
| **Nausea** | 1684 (84%) | 220 (11%) | 66 (3.3%) | 35 (1.7%) |
| **Headache** | 1135 (56.6%) | 539 (26.9%) | 195 (9.7%) | 136 (6.8%) |
| **Fatigue** | 1335 (66.6%) | 388 (19.4%) | 187 (9.3%) | 95 (4.7%) |
| **Muscle Pain/Myalgia** | 1235 (61.6%) | 439 (21.9%) | 217 (10.8%) | 114 (5.7%) |
| **Joint Pain** | 1463 (73%) | 336 (16.8%) | 129 (6.4%) | 77 (3.8%) |
| **Average** | 69.7% | 19.1% | 7.5% | 3.7% |

**Graph 6:**
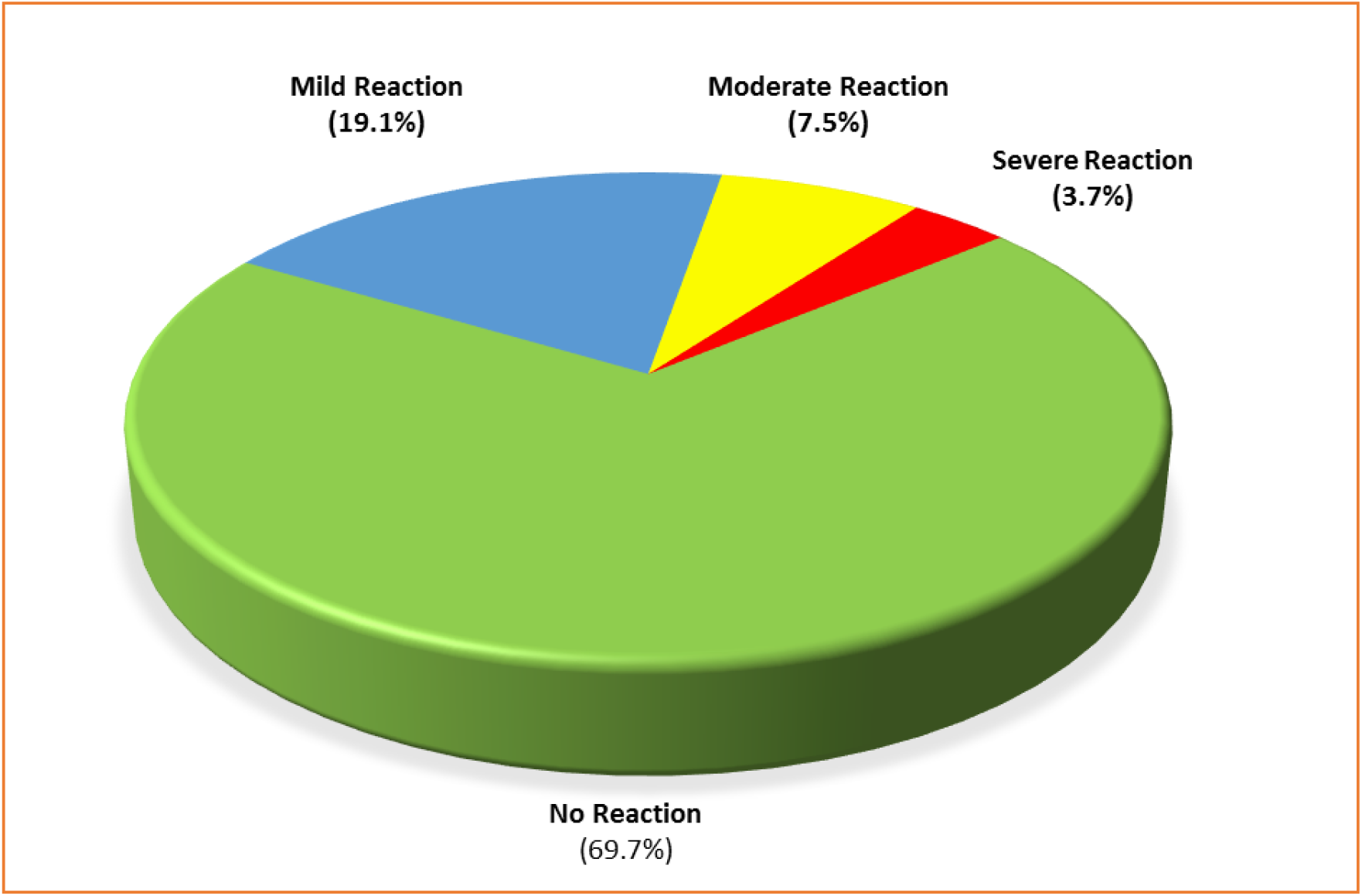
Intensity of commonly reported Adverse Reactions.

Combined for all commonly reported adverse reactions, the intensity was mild for 19.1%, moderate for 7.5% and severe for 3.7%. The severe adverse reactions reported were highest for fever (7.2%) followed by headache (6.8%), muscle pain/myalgia (5.7%) and pain at injection site (5.1%).

## Discussion

We set out to study the safety and antibody responses at baseline and 14±2 days following Covishield vaccine {ChAdOx1 nCoV-19 Corona Virus Vaccine (Recombinant)} in 1565 Health Care Workers. Seropositivity defined as SARS CoV-2 S1/S2 IgG titres>15 AU/ml is not a direct correlate of protection from SARS CoV-2 infection, however it can serve as a reasonable surrogate marker of protection (**11**).

In our study serostatus improved from 48.2% positive at baseline to 79.0% positive 2 weeks following first dose of vaccination. After first dose of vaccination overall higher percentage (98.2%) of seropositivity rates were observed in those with past history of COVID-19 disease **(Table no-9)** and heightened seroresponses **(Table no-6)** were seen in subgroup of subjects who were baseline seropositive compared to seronegative. From this observation it may be postulated that in those with past history of natural symptomatic/asymptomatic infection the prime dose may be acting as booster dose and such a group of individuals may not need a second dose. This early seroresponse data holds lot promise as it can have far reaching impact on vaccine availability for a larger population and thereby providing more widespread coverage. In a similar study by T Bradley et al it was observed that 3 weeks after a single vaccination, persons with recent SARS-CoV-2 infection or seropositive status had higher levels of antibody to four SARS-CoV-2 antigens and higher levels of antibodies with neutralizing characteristics than did those without a history of infection (**12**).

Like with roll out any new pharmaceutical product, safety of subject is of paramount importance and in this study we actively captured solicited local and systemic adverse events for 7 days post vaccination via daily diary record. Pain at injection site (62.7%) and soreness (24.1%) were most common solicited local adverse events, whereas fever (48.4%), headache (43.4%), myalgia (38.4%), fatigue (33.4%), joint pain (27.0%) and nausea (16.0%) were most common solicited systemic adverse events on day 1. Majority of local and systemic adverse events were seen in first 2 days post vaccination and thereafter the rates of adverse events dropped. Lesser reactogenicity was observed in subjects with age >50 years. None of the adverse events were severe enough to require hospitalization or emergency room visit. No major difference was observed in adverse events when subjects were stratified based on history of COVID-19 disease or baseline seropositivity. Similar observations were made by Rajeev Jayadevan^2^ (**13**). In a survey of symptoms following COVID-19 vaccination in India, in which 66 % of respondents reported at least one post-vaccination symptom. Tiredness (45%), myalgia (44%), fever (34%), headache (28%), local pain at injection site (27%), joint pain (12%), nausea (8%) and diarrhea (3%) were the most prevalent symptoms. All other symptoms were 1% or less. Vaccine associated Adverse Events of Special Interest (AESI), such as Vaccine Associated Enhanced Respiratory Disease (ERD) and any report of symptomatic COVID-19 were not reported in the first 14 days in our study.

## 4.0. Conclusion

After 1st vaccination as high as 79% of those vaccinated attained the positive antibody titers (SARS CoV-2 S1/S2 IgG titres>15 AU/ml). Among those with a past covid history, almost all attained these antibody titres. In comparison to baseline antibody titres, the magnitude of improvement has been of the order of 30 percentage points. The improvement observed was higher among relatively young and clinical HCW. Importantly there has been a mirror change in the values of IgG titre as evidenced from the fact that at the baseline about one-third had the lowest value of 3.8 whereas after 1st vaccination the same percentage had the highest value of more than 400.The common solicited adverse events reported were pain at the injection site, tenderness, fever, headache, muscle pain, fatigue, joint pain, and nausea. These symptoms are reported more by those with a history of covid and by those in the younger age group. The duration of the symptoms was 2 to 3 days and thereafter returned to normalcy. Vaccine associated Adverse Events of Special Interest (AESI), such as Vaccine Associated Enhanced Respiratory Disease (ERD) and any report of symptomatic COVID-19 were not reported in our study.

## Data Availability

Data is available with the Principal Investigator

## Acknowledgements

The investigators appreciate the support of Mr Manbir Singh, Senior Lab Technician, Microbiology, Neha Jakhar Trainee Microbiology; the Clinical Research team consisting of Mr Dheeraj Sharma, Ms Rita Maity, Mr Rehan Khan, Mr Bajarang, Ms Arunima, Ms Ruchika, Mr Iqbal, Ms Rhea Aggarwal, Clinical research and Pharmacy interns from Apeejay Stya University, GD Goenka University, Jamia Hamdard University, MGIMS Sewagram, Mr Rajiv Sikka and the IT team at Medanta, Mr Pankaj Sahni (CEO) and all staff of the Medanta-The Medicity Hospital, Gurugram and all the participants of the research project.

Editorial support in manuscript writing by Mr Radhesh Notiyal

Funding support from an intramural grant of the Medanta Institute of Education and Research

